# Gut microbiome compositional and functional features associate with Alzheimer’s disease pathology

**DOI:** 10.1101/2024.09.04.24313004

**Authors:** Jea Woo Kang, Lora A. Khatib, Margo B. Heston, Amanda H. Dilmore, Jennifer S. Labus, Yuetiva Deming, Leyla Schimmel, Colette Blach, Daniel McDonald, Antonio Gonzalez, MacKenzie Bryant, Karenina Sanders, Lara Schwartz, Tyler K. Ulland, Sterling C. Johnson, Sanjay Asthana, Cynthia M. Carlsson, Nathaniel A. Chin, Kaj Blennow, Henrik Zetterberg, Federico E. Rey, Alzheimer Gut Microbiome Project Consortium, Rima Kaddurah-Daouk, Rob Knight, Barbara B. Bendlin

**Author notes:** These three authors contributed equally to this work (Jea Woo Kang, Lora A. Khatib, Margo B. Heston).

## Abstract

**BACKGROUND:** The gut microbiome is a potentially modifiable factor in Alzheimer’s disease (AD); however, understanding of its composition and function regarding AD pathology is limited.

**METHODS:** Shallow-shotgun metagenomic data was used to analyze fecal microbiome from participants enrolled in the Wisconsin Microbiome in Alzheimer’s Risk Study, leveraging clinical data and cerebrospinal fluid (CSF) biomarkers. Differential abundance and ordinary least squares regression analyses were performed to find differentially abundant gut microbiome features and their associations with CSF biomarkers of AD and related pathologies.

**RESULTS:** Gut microbiome composition and function differed between people with AD and cognitively unimpaired individuals. The compositional difference was replicated in an independent cohort. Differentially abundant gut microbiome features were associated with CSF biomarkers of AD and related pathologies.

**DISCUSSION:** These findings enhance our understanding of alterations in gut microbial composition and function in AD, and suggest that gut microbes and their pathways are linked to AD pathology.

## 1 BACKGROUND

The human gut microbiome is recognized as an important modifiable factor in health and disease. It is related to overall gut health by maintaining gut barrier integrity and gut immune homeostasis via balanced composition and production of microbial metabolites such as short-chain fatty acids (SCFAs).^1–3^ However, in certain disease states, including Alzheimer’s disease (AD), gut microbiome composition and its metabolic changes may alter and exacerbate the disease.

Compositional differences in gut microbiota, including relative abundance and diversity, have been observed between control and AD groups.^4^ Other studies have reported that gut microbiome composition is altered among people with AD dementia, individuals with mild cognitive impairment (MCI), or preclinical AD compared with healthy controls.^5–8^ To better determine the relationship between gut microbiome and AD pathology, studies have leveraged measures of AD biomarkers obtained via cerebrospinal fluid (CSF) analysis,^9^ positron emission tomography (PET),^10^ and plasma.^11^ Additionally, inflammatory markers have been utilized to find associations with gut inflammation-driven AD pathology.^12,13^

The shift towards functional analysis provides deeper insights into the functional potential of the microbiome, which is important given that multiple lines of evidence indicate that gut microbial pathways and associated metabolites influence disease development, as well as providing the opportunity to identify therapeutic targets.^14–17^ A small number of studies have examined gut microbiome composition and function together with markers of AD pathology in humans,^8,18–20^ but many other studies to date are limited to the composition of gut microbes in a single cohort. The Alzheimer Gut Microbiome Project (AGMP) initiative continues to leverage gut microbiome and metabolome to better understand metabolic processes that influence AD pathology.

To identify differences in composition and function as well as relationships between gut microbiome and AD pathology, this study collected stool samples from participants enrolled in the Microbiome in Alzheimer’s Risk Study (MARS). Differences in microbiome diversity as well as abundance were compared between AD-related groups (diagnosis, amyloid status, and *APOE* ε*4* status), and the co-occurrence of the common gut microbiota features was analyzed among comparison groups. In addition, validation of gut microbiota composition that was differentially abundant between AD compared to healthy controls was performed in a larger cohort of participants who are part of the AGMP. Furthermore, determination of microbiome functional differences was compared between people with AD dementia and cognitively unimpaired (CU) individuals. Lastly, associations between any differentially abundant gut microbiome features were examined in relation to CSF biomarkers of AD and related pathologies to identify the potential microbes and microbial pathways that relate to AD pathology. We hypothesized that gut microbiome alterations in composition and function would be present among people with AD dementia compared to CU, as well as associate with AD pathology.

## 2 METHODS

### 2.1 Participants

Participants included in this study were recruited from the Wisconsin Alzheimer’s Disease Research Center (ADRC) Clinical Core and the Wisconsin Registry for Alzheimer’s Prevention (WRAP).^21^ The WRAP study enrolled participants between the ages of 40–65 years at study entry, and the cohort is enriched for parental history of AD dementia. The Wisconsin ADRC clinical core enrolls participants who span the clinical and biological spectrum of AD, from those who are CU to individuals with mild cognitive impairment (MCI) and AD dementia. Participants underwent *APOE* genotyping using competitive allele-specific PCR-based KASP™ genotyping assays (LGC Genomics, Beverly, MA)^22^ as well as longitudinal assessments of cognition and laboratory tests. Biomarkers of AD determined with CSF collection and PET neuroimaging were collected in a subset of the cohort. Participants underwent fecal sample collection as part of their participation in MARS, which was used to analyze the gut microbiome. Participants completed questionnaires including medical history and diet, at the time of fecal sample collection.

Participant diagnosis of AD was determined by a multidisciplinary consensus diagnostic panel and based on the National Institute on Aging–Alzheimer’s Association (NIA-AA) criteria.^23,24^ Participants underwent dynamic [C-11]Pittsburgh compound B (PiB) PET scans and lumbar puncture for CSF collection to determine their amyloid status. Amyloid positivity on PET imaging was achieved by the visual rating (1.19 or greater)^25^ from a global PiB distribution volume ratio (DVR) and determined for CSF via the Aβ_42_/Aβ_40_ ratio (less than 0.046).^26^ The study procedures were approved by the University of Wisconsin Institutional Review Board, and all participants provided signed or oral informed consent.

An independent cohort of participants who are part of the AGMP and who were recruited from multiple NIA-funded ADRC across the U.S. (n = 448, **Table S1**) was included for validation of differential abundance analysis. AGMP participants from Wisconsin were excluded to ensure a unique validation sample.

### 2.2 Fecal sample collection and metagenomic data sequencing

Fecal samples were collected as previously described.^4^ Briefly, participants collected their stool samples at home with provided fecal collection kits. Participants returned their samples in insulated containers which were chilled with frozen gel packs. Returned samples were immediately weighed and scored on the Bristol stool scale. Fecal samples were then subsampled using sterile straws and stored at −80°C until processing.

Fecal samples were processed for DNA extraction as previously described.^27^ Briefly, samples were extracted using the MoBio PowerMag Soil DNA isolation kit with a magnetic bead plate. Extracted genomic DNA (gDNA) was quantified with Quant-iT PicoGreen double-stranded DNA (dsDNA) assay kit (Thermo Fisher Scientific Inc.), and underwent a miniaturized KAPA HyperPlus library preparation using an iTru indexing strategy.^28,29^ Library was normalized pooled based on concentration, PCR cleaned, and then size selected (300-700 bp) on the Sage Science PippinHT. Libraries were sequenced on an Illumina NovaSeq 6000 as a paired-end 150-cycle run at the University of California San Diego (UCSD) IGM Genomics Center as part of AGMP initiative.^30^

### 2.3 Metagenomic data processing

The metagenomic data processing was performed as previously described.^31^ The sequence data were filtered for all adapters known to fastp (version 0.23.4) in paired-end mode by explicitly specifying a known adapters file.^32^ Fastp also removed sequences shorter than 45 nucleotides with –l, a flag to filter the minimum length of each sequence. Each sample was then filtered against each genome in the human pangenome,^33^ as well as both T2T-CHM13v2.0^34^ and GRCh38,^35^ using minimap2^36^ (version 2.26-r1175) with “-ax sr” for short read mode. The data were first run in paired-end mode, and then run in single-end mode, per genome. Each successive run was converted from SAM to FASTQ using samtools^37^ (version 1.17) with arguments –f 12 –F 256 –N for paired-end data and –f 4 –F 256 for single-end. The single-end data are repaired using fastq_pair^38^ (version 1.0) specifying a table size of 50M with –t. Compute support was provided with GNU Parallel^39^ (version 20180222). Single-end FASTQ output from samtools was split into R1 and R2 with a custom Rust program, with rust-bio for parsing^40^ (version 1.4.0). Data were multiplexed with sed and demultiplexed using a custom Python script. Shotgun sequencing data were then uploaded to and processed through Qiita^41^ (Study ID 13663). Sequence adapter and host filtering were executed using qp-fastp-minimap2 version 2022.04. Subsequently, Woltka^42^ version 0.1.4 (qp-woltka 2022.09) with the Web of Life 2 database was employed for taxonomic and functional predictions. Genomic coverages were computed, and features with less than 25% coverage were excluded.^43^ To enhance data quality, a prevalence filter using QIIME 2 v2023.5^44^ was applied, eliminating features present in less than 10% of samples and samples with a sampling depth of less than 500,000 reads to mitigate the inclusion of erroneous and low-quality reads. The resulting feature table was utilized for downstream analysis.

### 2.4 Biomarker measurements

#### 2.4.1 CSF biomarkers

CSF samples were collected via lumbar puncture in the morning after fasting for 8-12 hours as previously described.^26^ CSF biomarkers were measured using the NeuroToolKit (NTK), a panel of exploratory robust prototype assays (Roche Diagnostics International Ltd, Rotkreuz, Switzerland). The following biomarkers were quantified on the Cobas^®^ e 601 module (Roche Diagnostics International Ltd, Rotkreuz, Switzerland): Aβ_42_, pTau_181_, tTau, S100 calcium binding protein B (S100B), and interleukin-6 (IL-6), and the remaining biomarkers were assayed on the Cobas^®^ e 411 analyzer: Aβ_40_, neurofilament light protein (NfL), neurogranin, α-synuclein, glial fibrillary acidic protein (GFAP), chitinase-3-like protein 1 (YKL-40), and soluble triggering receptor expressed on myeloid cells 2 (sTREM2). Each biomarker was measured as markers of AD and related pathologies which were amyloid pathology (Aβ_42_/Aβ_40_), tau pathophysiology (pTau_181_ and tTau), neurodegeneration (NfL), synaptic dysfunction and injury (neurogranin and α-synuclein), inflammation (IL-6), and glial activation (S100B, GFAP, YKL-40, and sTREM2).

#### 2.4.2 PiB PET biomarker

Dynamic ^11^C-PiB scans were acquired using a Siemens ECAT EXACT HR+ tomograph as previously described.^45^ DVRs were estimated with Logan graphical analysis and a threshold of 1.19 for global DVR was used to determine PiB status which was used along with CSF Aβ_42_/Aβ_40_ ratio to confirm amyloid status of participants.

### 2.5 Statistical analysis

Statistical analysis on participant demographics was performed across clinical diagnoses using the Kruskal-Wallis rank sum test for continuous variables and Pearson’s Chi-squared test for categorical variables. Analysis with multiple comparisons was corrected for multiple tests employing the Bonferroni correction method. Gut microbiome diversities were calculated using QIIME 2 tools.^44^ Alpha diversity indices were calculated including Shannon,^46^ Evenness,^47^ and Faith’s phylogenetic diversity (PD).^48^ Beta diversity indices were calculated including Bray– Curtis dissimilarity,^49^ and Weighted^50^ and Unweighted^51^ UniFrac. The Bayesian Inferential Regression for Differential Microbiome Analysis (BIRDMAn) pipeline was used for microbiome differential abundance (DA) analysis.^52^ Microbiome features (composition and function) were outcome variables and each AD-related group (clinical diagnosis, amyloid status, and *APOE* ε*4* status) was a predictor adjusting for covariates including age, sex, BMI, Bristol type, medication status, and age difference between fecal collection and measurements of each predictor variable. Log ratios of Top and Bottom features of each AD-related group (log[Top features/Bottom features]) were calculated and analyzed using the Mann–Whitney *U* test to identify similarities in microbiome features that differ between AD-related groups. Venn diagrams were created to illustrate any overlapping taxonomies across AD groups in each Top (more abundant in AD-related groups) and Bottom (less abundant in AD-related groups) group at each taxonomic level (phylum, family, genus, and species). Log ratios of Top and Bottom features of each AD group (log[Top features/Bottom features]) were also calculated and analyzed using the Mann–Whitney *U* test in MARS and validation cohorts to identify similarities in microbiome features that differ between AD and CU groups in both cohorts. Log ratios of Top and Bottom functional features (log[Top features/Bottom features]) were calculated and statistical significance was determined by the Mann–Whitney *U* test between AD and CU groups. Differentially abundant KEGG Orthology (KO)^53^ pathways and their associated species were determined using BIRDMAn.^52^ Robust Aitchison principal component analysis (RPCA) from Gemelli (version 0.0.10) was used to analyze sparse compositional KO^53^ microbiome pathway features that are separated by sample variations.^54^ RPCA results were visualized with scores plots and biplots. Statistical analysis on RPCA was performed with permutational multivariate analysis of variance (PERMANOVA)^55^ between groups.

To explore the relationships between key microbial features and pathways identified via BIRDMAn and CSF biomarkers of AD and related pathologies, we applied an ordinary least squares (OLS) linear regression approach. Prior to fitting the linear regression model, CSF biomarkers were standardized using the StandardScaler (version 0.24.1) from the scikit-learn library.^56^ To address issues with sparse, compositional data, we used the multiplicative_replacement function from the scikit-bio (version 0.5.7) skbio.stats.composition module to preprocess the metagenomics data. This function replaces zeros with small positive values, preserving the compositional nature of the data. Subsequently, a centered log ratio (CLR) transformation was applied to the metagenomics data to account for compositionality. Finally, ordinary least squares (OLS) linear regression was performed on the microbial features differentially abundant in AD versus CU, in relation to each CSF biomarker. The results were visualized using heatmaps.

All other statistical analyses were performed using Python libraries SciPy (1.13.0),^57^ scikit-learn (1.4.2),^56^ and NumPy (version 1.26.4).^58^ All figures were generated using Python libraries Matplotlib (version 3.6.0)^59^ and Seaborn (version 0.11.2).^60^

## 3 RESULTS

### 3.1 Participant demographics

Participant characteristics are shown by clinical diagnosis in **Table 1**. Participants were aged between 47-93 years. The mean age differed significantly in dementia-AD vs CU. The percent ratio of *APOE* ε*3/*ε*3* carriers was significantly lower and the percent ratio of *APOE* ε*4/*ε*4* carriers was higher in dementia-AD compared to CU. Amyloid positivity was higher in the AD dementia group.

**Table 1.**
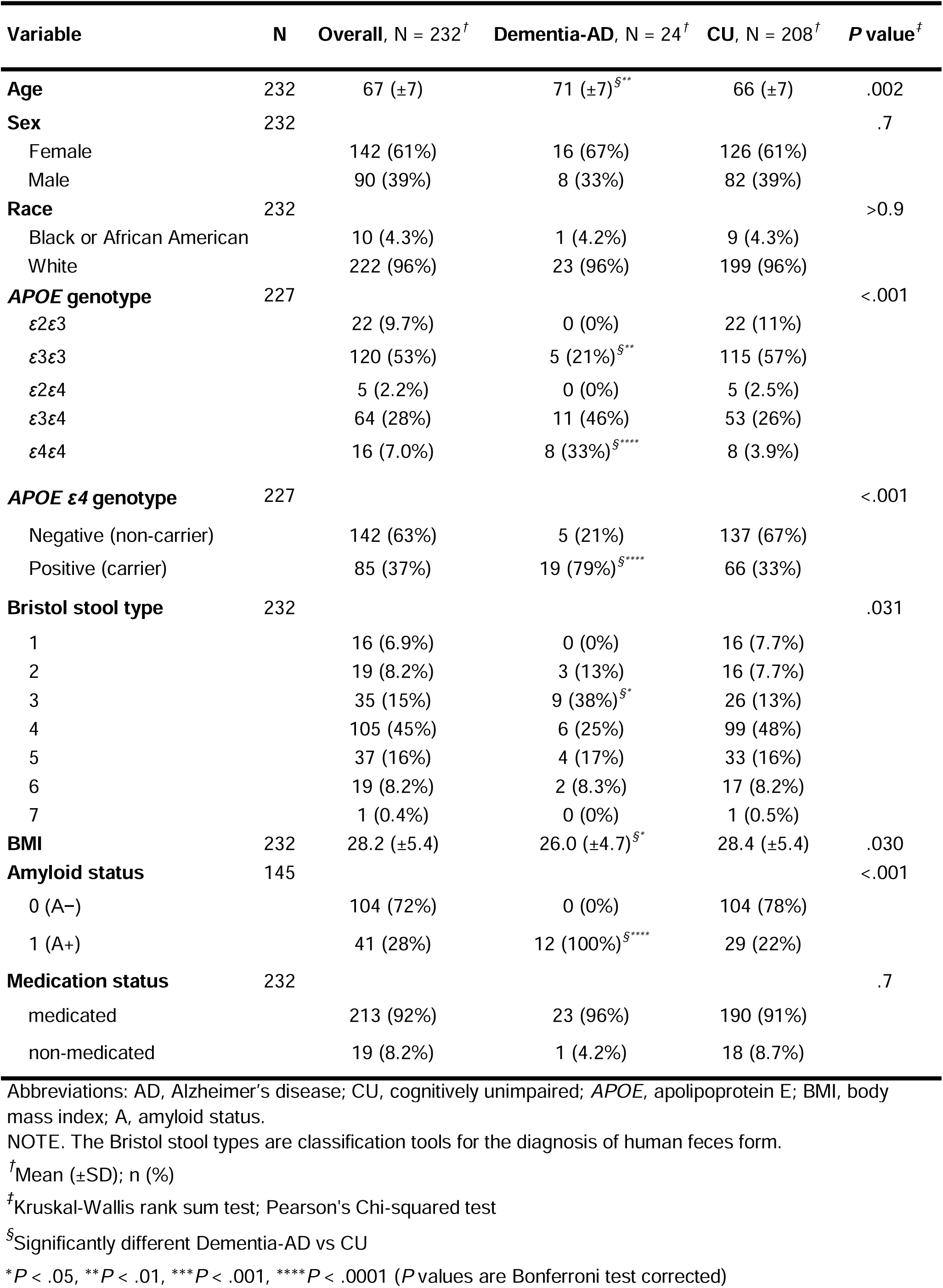
Participant demographics at fecal sample collection by clinical diagnosis.

### 3.2 Diversity results in gut microbiota composition

Alpha diversity indices (Shannon, Evenness, and Faith’s PD) were presented on the y-axis and all AD group categories were presented on the x-axis (**Figure 1A-I**). The only comparison with significant alpha diversity differences was the *APOE* ε*4* comparison (**Figure 1G and H**). All alpha diversity indices showed significant differences or a trend toward differences between *APOE* ε*4*+ and *APOE* ε*4*− groups. Individuals who were *APOE* ε*4*− had significantly higher alpha diversity than *APOE* ε*4+*.

**Figure 1.**
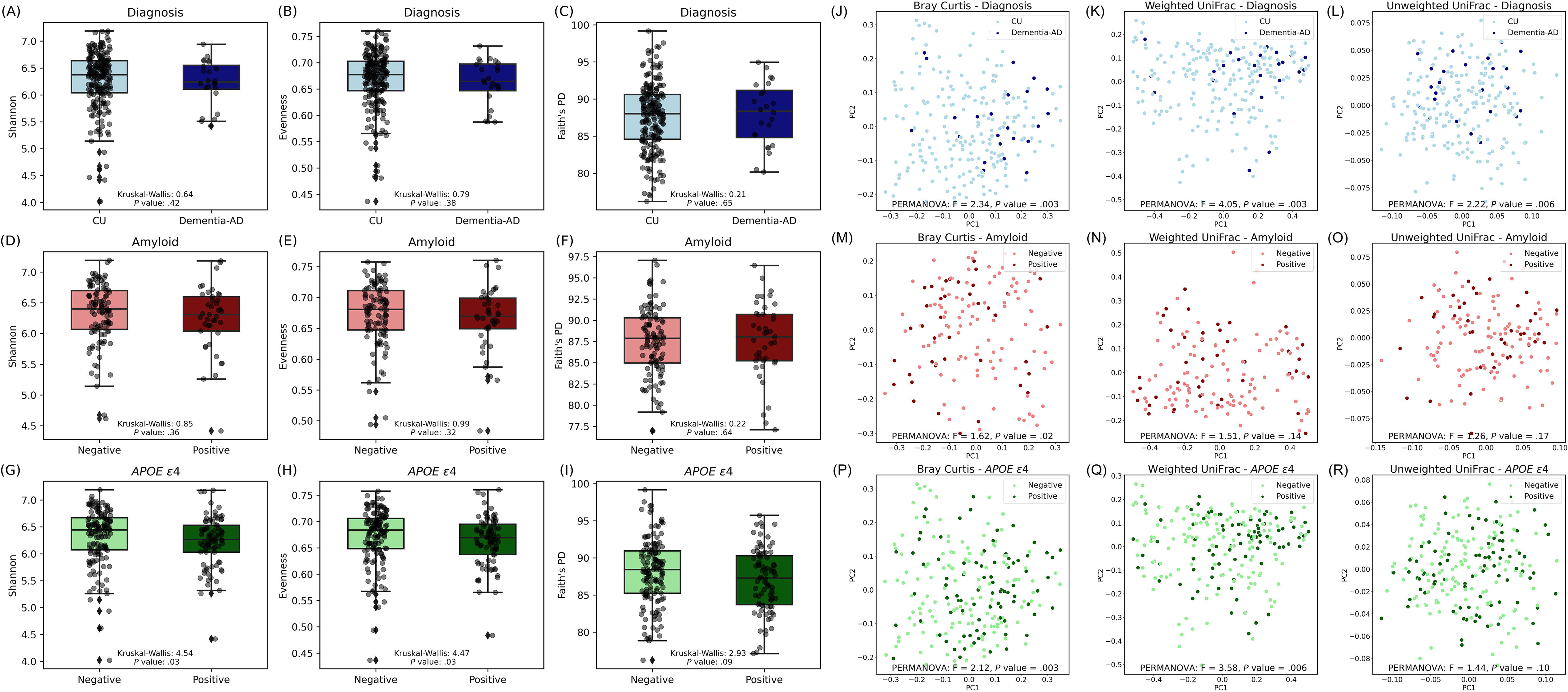
Alpha and beta diversity metrics across AD groups. (A-C) Shannon, Evenness, and Faith’s PD metrics for individuals categorized by clinical diagnosis (CU vs. Dementia-AD). (D-F) Metrics for amyloid status (Negative vs. Positive). (G-I) Metrics across *APOE* ε*4* status (Negative vs. Positive). Each box plot is overlaid with individual data points, enhancing visualization of the data distribution within each group. Kruskal-Wallis test was used to determine statistical significance. (J-L) Differences in beta diversity metrics (Bray Curtis, Weighted UniFrac, and Unweighted UniFrac, respectively) for individuals categorized by clinical diagnosis (CU vs. Dementia-AD). (M-O) Metrics for amyloid status (Negative vs. Positive). (P-R) Metrics across *APOE* ε*4* status (Negative vs. Positive). Principal coordinates (PC)1 and PC2 axes represent the most variance in data. Each plot is color-coded by the respective group, highlighting the spatial distribution and clustering based on the dissimilarity indices. PERMANOVA was used to determine statistical significance.

Beta diversity indices (Bray–Curtis dissimilarity, and Weighted and Unweighted UniFrac) were visualized with principal coordinates analysis (PCoA) plots for each AD group (**Figure 1J-R**). Significant differences between groups based on clinical diagnosis (diagnosis) were observed with each metric (**Figure 1J-L**). Differences in beta diversity between amyloid-positive and amyloid-negative individuals were detected only with the Bray–Curtis dissimilarity index (**Figure 1M-O**). Individuals positive and negative for *APOE* ε*4* demonstrated differences with both the Bray-Curtis and Weighted UniFrac metrics, but not with Unweighted UniFrac (**Figure 1P-R**), suggesting that the relative abundance of major taxa rather than community membership is important in driving these differences.

### 3.3 Gut microbiota composition in clinical diagnosis, amyloid status, and *APOE* ε*4* status groups

A DA analysis on gut microbiota composition was performed based on clinical diagnosis, amyloid status, and *APOE* ε*4* status groups using BIRDMAn and visualized as forest plots (**Figure 2**). Gut microbiota taxonomic features that showed the most differences by the effect size (log ratio) in each comparison group were displayed up to 20 features in each Top (more abundant) and Bottom (less abundant) group for taxonomic levels including phylum, family, genus, and species. DA analysis between AD and CU in clinical diagnosis showed distinct gut microbiota composition at each taxonomic level (**Figure 2A, D, G, and J**). At the phylum level, the abundance of phylum Firmicutes_A was lower in AD compared to CU (Bottom), and the abundance of phyla Bacteroidota, Patescibacteria, and Fusobacteriota was higher in AD compared to CU (Top) (**Figure 2A**). At the family level, families such as *Clostridiaceae*, *Turicibacteraceae*, *Pasteurellaceae*, *Dialisteraceae*, *Enterococcaceae*, and *Ruminococcaceae* were in the Bottom group and *Fusobacteriaceae*, *Nanogingivalaceae*, *Gemellaceae*, and *Bacteroidaceae* were in the Top group (**Figure 2D**). At the genus level, genera including *Clostridium_P*, *Ruminococcus*, and *Cryptobacteroides* were in the Bottom group and *Fusobacterium_A*, *Fusobacterium*, *Nanogingivalis*, and *Gemella* were in the Top group (**Figure 2G**). At the species level, species included in the Bottom group were *Cryptobacteroides* spp., *Clostridium_P perfringens*, *Turicibacter sanguinis*, *Prevotella hominis*, and *Prevotella copri*, and in the Top group were *Fusobacterium_A mortiferum*, *Fusobacterium nucleatum*, *Fusobacterium animalis*, *Nanogingivalis gingivitcus*, *Collinsella stercoris*, and *Collinsella tanakaei* (**Figure 2J**).

**Figure 2.**
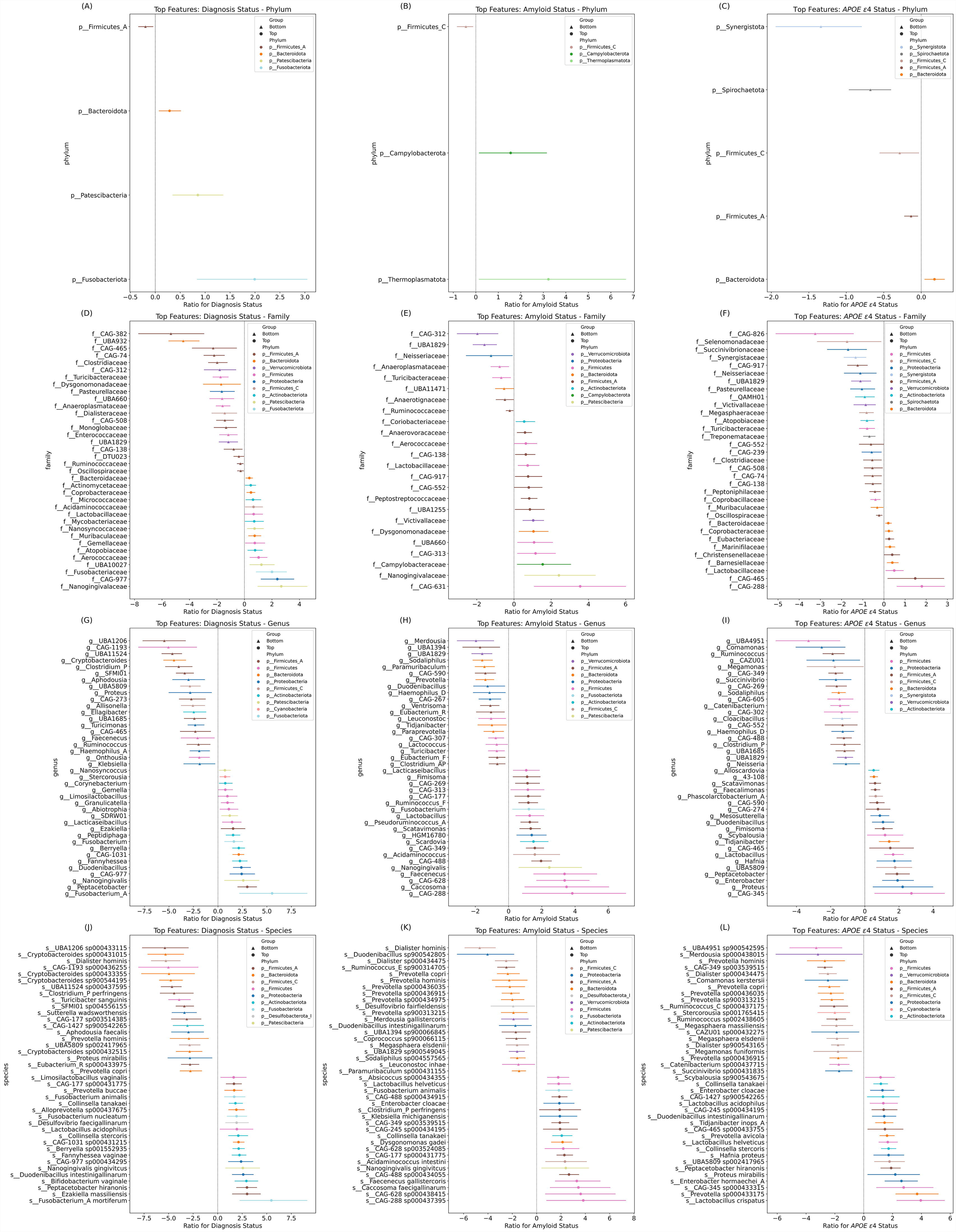
Differential abundance (DA) across AD groups. Forest plots illustrating the DA of microbial features associated with AD groups. (A, D, G, and J) Contrasts in the abundance of various bacterial taxa at (A) phylum, (D) family, (G) genus, and (J) species levels between AD dementia and CU. (B, E, H, and K) The differences in abundance at these taxonomic levels between A+ and A− individuals. (C, F, I, and L) The microbial features differentially abundant between *APOE* ε*4*+ and *APOE* ε*4*− groups. The x-axes quantify the log ratio of presence between groups, with values above one indicating a higher abundance in the first-mentioned group. Circles denote “Top” features, indicating a positive association with AD groups (dementia diagnosis, amyloid positivity, and *APOE* ε*4* positivity), whereas triangles denote “Bottom” features, indicating a negative association. The lines are color-coded by unique phylum as labeled in the legend. DA analysis was conducted using BIRDMAn.

DA analysis between A+ and A− in amyloid status showed distinct gut microbiota composition at each taxonomic level (**Figure 2B, E, H, and K**). At the phylum level, the abundance of phylum Firmicutes_C was lower in A+ compared to A− (Bottom), and the abundance of phyla Thermoplasmatota and Campylobacterota was higher in A+ compared to A− (Top) (**Figure 2B**). At the family level, families such as *Neisseriaceae*, *Anaeroplasmataceae*, *Turicibacteraceae*, and *Ruminococcaceae* were in the Bottom group and *Nanogingivalaceae*, *Campylobacteraceae*, and *Coriobacteriaceae* were in the Top group (**Figure 2E**). At the genus level, genera including *Prevotella*, *Eubacterium_R*, and *Lactococcus* were in the Bottom group and *Fusobacterium*, *Nanogingivalis*, and *Acidaminococcus* were in the Top group (**Figure 2H**). At the species level, species included in the Bottom group were *Dialister hominis*, *Prevotella copri*, and *Prevotella hominis*, and in the Top group were *Nanogingivalis gingivitcus*, *Collinsella tanakaei*, and *Fusobacterium animalis* (**Figure 2K**).

DA analysis between *APOE* ε*4*+ and *APOE* ε*4*− in *APOE* ε*4* status analyses showed distinct gut microbiota composition at each taxonomic level (**Figure 2C, F, I, and L**). At the phylum level, the abundance of phyla Firmicutes_A, Firmicutes_C, Spirochaetota, and Synergistota was lower in *APOE* ε*4*+ compared to *APOE* ε*4*− (Bottom), and the abundance of phylum Bacteroidota was higher in *APOE* ε*4*+ compared to *APOE* ε*4*− (Top) (**Figure 2C**). At the family level, families such as *Selenomonadaceae*, *Neisseriaceae*, *Pasteurellaceae*, *Turicibacteraceae*, and *Clostridiaceae* were in the Bottom group and *Lactobacillaceae*, *Eubacteriaceae*, and *Bacteroidaceae* were in the Top group (**Figure 2F**). At the genus level, genera including *Ruminococcus* and *Clostridium_P* were in the Bottom group and *Enterobacter*, *Hafnia*, and *Lactobacillus* were in the Top group (**Figure 2I**). At the species level, species included in the Bottom group were *Prevotella hominis*, *Dialister* spp., and *Ruminococcus* spp., and in the Top group were *Enterobacter hormaechei_A*, *Hafnia proteus*, *Collinsella stercoris*, *Enterobacter cloacae*, and *Collinsella tanakaei* (**Figure 2L**).

Log ratios of microbiome counts were calculated between the sum of the Top and Bottom groups from DA analysis to test the overall significant differences of microbiome features between AD conditions (log[sum of Top features/sum of Bottom features]) (**Figure 3**). Overall, there were significant differences in each AD-related group (diagnosis, amyloid, and *APOE* ε*4*) (**Figure 3A, E, and I**). Features that significantly differed between AD and CU also differed between *APOE* ε*4*+ and *APOE* ε*4*− (**Figure 3A and C**). Features that significantly differed between A+ and A− also differed between AD and CU (**Figure 3D and E**). Features that significantly differed between *APOE* ε*4*+ and *APOE* ε*4*− also differed between AD and CU (**Figure 3G and I**).

**Figure 3.**
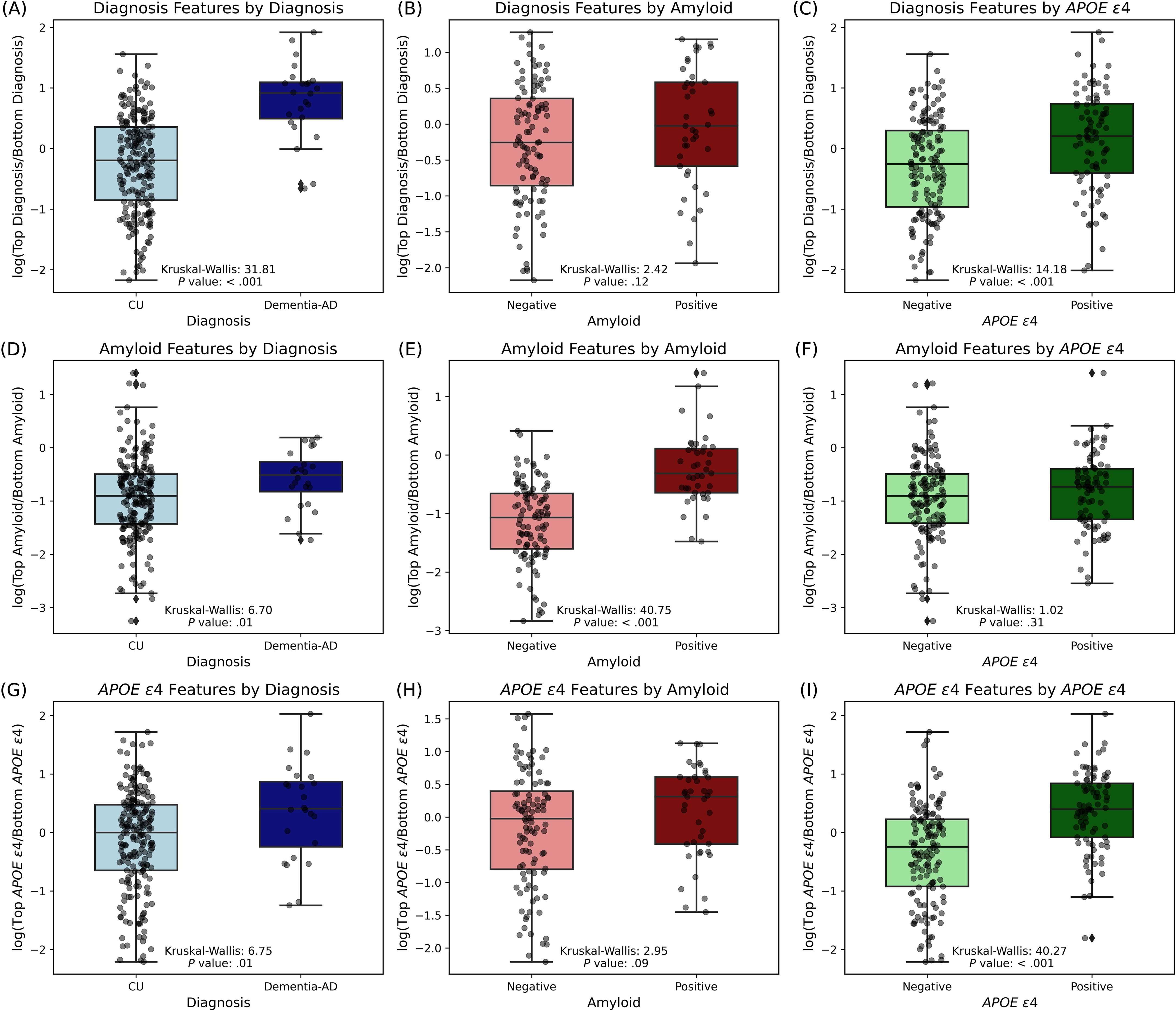
Comparative analysis of top and bottom features across AD groups. Box plots comparing the distribution of log-transformed ratios of differentially abundant microbial species in relation to diagnosis, amyloid status, and *APOE* ε*4* status. (A-C) The log-transformed ratios of microbes for diagnosis groups (AD vs CU). (D-F) The log-transformed ratios of amyloid-related microbes (A+ vs A−). (G-I) The log-transformed ratios of *APOE* ε*4*-related microbes (*APOE* ε*4*+ vs *APOE* ε*4*−). Each column compares the CU and AD groups (A, D, and G), A+ and A− groups (B, E, and H), and *APOE* ε*4*+ and *APOE* ε*4*− groups (C, F, and I). Each panel includes a Kruskal-Wallis test statistic and associated *P* value, indicating the statistical significance of the differences observed.

### 3.4 Common gut microbiota features in clinical diagnosis, amyloid status, and *APOE* ε*4* status groups

Venn diagrams were used to find common microbiome features across different AD conditions for each Bottom and Top group at each taxonomic rank (**Table S2**). At the phylum level, phyla Bacteroidota co-occurred between diagnosis and *APOE* ε*4* in the Top group (**Figure 4A**). Firmicutes_A co-occurred between diagnosis and *APOE* ε*4*, and Firmicutes_C co-occurred between amyloid and *APOE* ε*4* in the Bottom group (**Figure 4B**). In the Top group at the family level, the family *Lactobacillaceae* co-occurred across all conditions, families *Bacteroidaceae* and *Coprobacteraceae* between diagnosis and *APOE* ε*4*, and families *Nanogingivalaceae* and *Aerococcaceae* co-occurred between diagnosis and amyloid (**Figure 4C**). In the Bottom group, *UBA1829* and *Turicibacteraceae* co-occurred across all conditions (diagnosis, amyloid, and *APOE* ε*4*), families *CAG-508*, *CAG-74*, *Oscillospiraceae*, *Clostridiaceae*, *Pasteurellaceae*, and *CAG-138* co-occurred between diagnosis and *APOE* ε*4*, families *Ruminococcaceae*, *Anaeroplasmataceae*, and *CAG-312* co-occurred between diagnosis and amyloid, and family *Neisseriaceae* co-occurred between amyloid and *APOE* ε*4* (**Figure 4D**). Multiple genera and species co-occurred across all conditions (**Figure 4E-H**). In the Top group, the genus *Veillonella_A* co-occurred across all conditions, and the following numbers of genera co-occurred between each intersection, i.e., diagnosis and *APOE* ε*4*: 7, diagnosis and amyloid: 6, and amyloid and *APOE* ε*4*: 4 (**Figure 4E, Table S2**). In the Bottom group, 13 genera co-occurred across all conditions including *Prevotella* and *Turicibacter*, and the following numbers of genera co-occurred between each intersection, i.e., diagnosis and *APOE* ε*4*: 30, diagnosis and amyloid: 9, and amyloid and *APOE* ε*4*: 8 (**Figure 4F, Table S2**). In the Top group at the species level, 12 species including *Bacteroides ovatus*, *Collinsella tanakaei*, *Prevotella corporis*, and more co-occurred across all conditions, and the following numbers of species co-occurred between each intersection, i.e., diagnosis and *APOE* ε*4*: 18, diagnosis and amyloid: 10, and amyloid and *APOE* ε*4*: 7 (**Figure 4G, Table S2**). In the Bottom group, 27 species including *Ruminococcus_C callidus*, *Dialister succinatiphilus*, *Prevotella copri*, and more co-occurred across all conditions, and the following numbers of species co-occurred between each intersection, i.e., diagnosis and *APOE* ε*4*: 42, diagnosis and amyloid: 13, and amyloid and *APOE* ε*4*: 16 (**Figure 4H, Table S2**).

**Figure 4.**
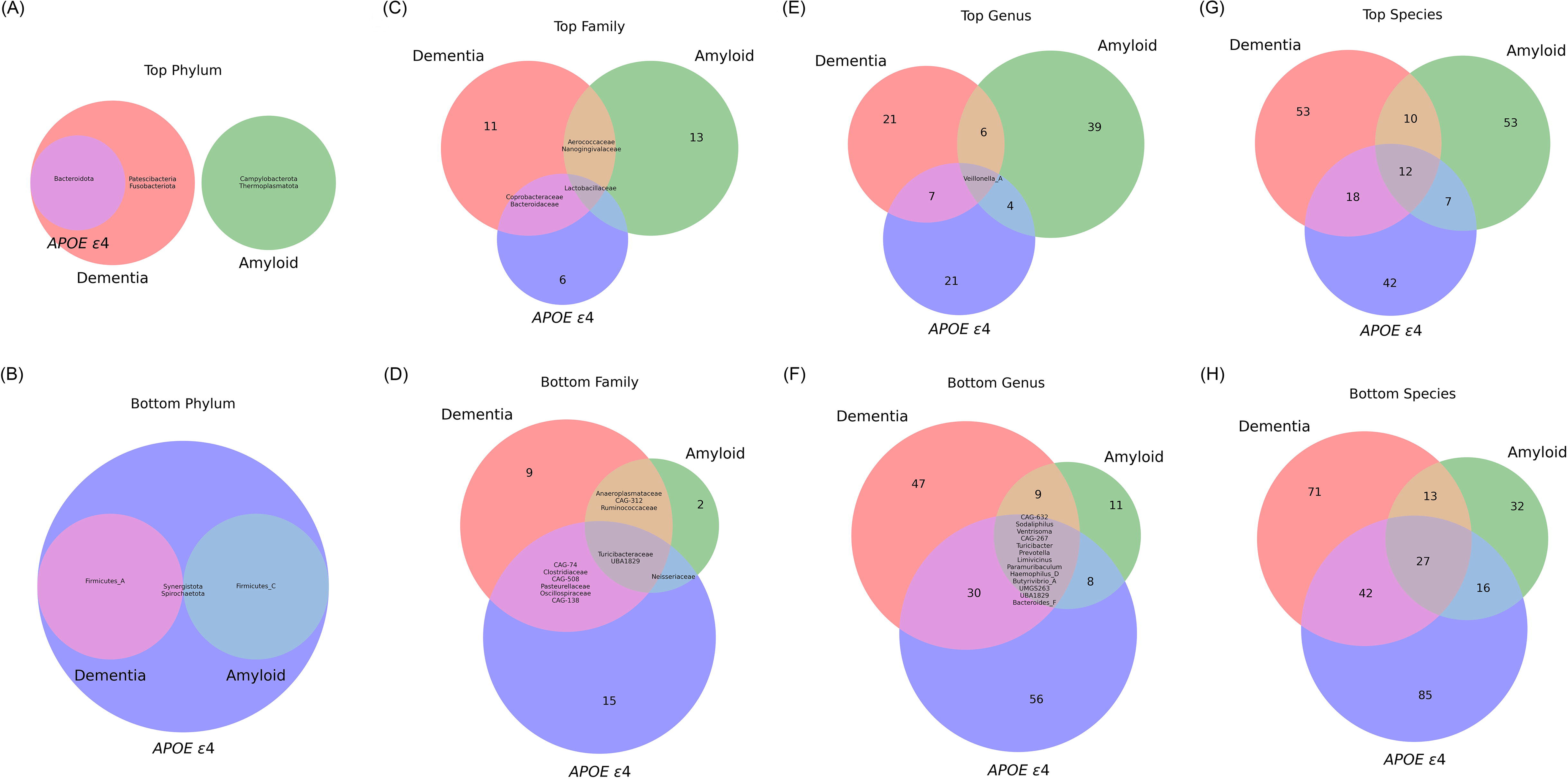
Venn-diagram of co-occurrence of microbial features across AD groups. The diagrams on the left column (A, C, E, and G) depict the Top (positively-associated) differentially abundant features, while those on the right column (B, D, F, and H) show the Bottom (negatively-associated) differentially abundant features. (A and B) Top and Bottom microbial phyla, respectively. These diagrams identify unique and shared phyla associated with each of the three AD groups. (C and D) Top and Bottom microbial families, respectively. These diagrams highlight the family-level microbial differences that correlate with AD diagnosis, amyloid presence, and *APOE* ε*4* genotype presence. (E and F) Top and Bottom microbial genera, respectively. These diagrams provide insight into the genus-level microbial composition influenced by the specified AD groups. (G and H) Top and Bottom microbial species, respectively. These diagrams detail the number of species that are unique and shared across the three AD groups. Each diagram contains colored regions representing intersections between the groups: red for dementia, green for amyloid, and blue for *APOE* ε*4*. The numbers within each segment of the diagrams indicate the count of microbial features unique to or shared between the conditions. Specific microbial features are listed in **Table S2**.

### 3.5 Validation of shallow-shotgun data with the ADRC dataset on gut microbiota composition

To validate the features linked with dementia-AD (AD) compared to healthy controls (CU), we tested the log-transformed ratios of features more and less abundant in AD in the MARS cohort against a larger cohort of participants who are part of the AGMP and who were recruited from multiple NIA-funded ADRC across the U.S. (n = 448; **Figure 5**). AGMP participants from Wisconsin were excluded to ensure a unique validation sample. The features that were differentially abundant in the MARS cohort (Kruskal-Wallis: 31.81, *P* value: < .001; **Figure 5A**) were also found to be differentially abundant in the larger validation cohort (Kruskal-Wallis: 5.59, *P* value: .02; **Figure 5B**).

**Figure 5.**
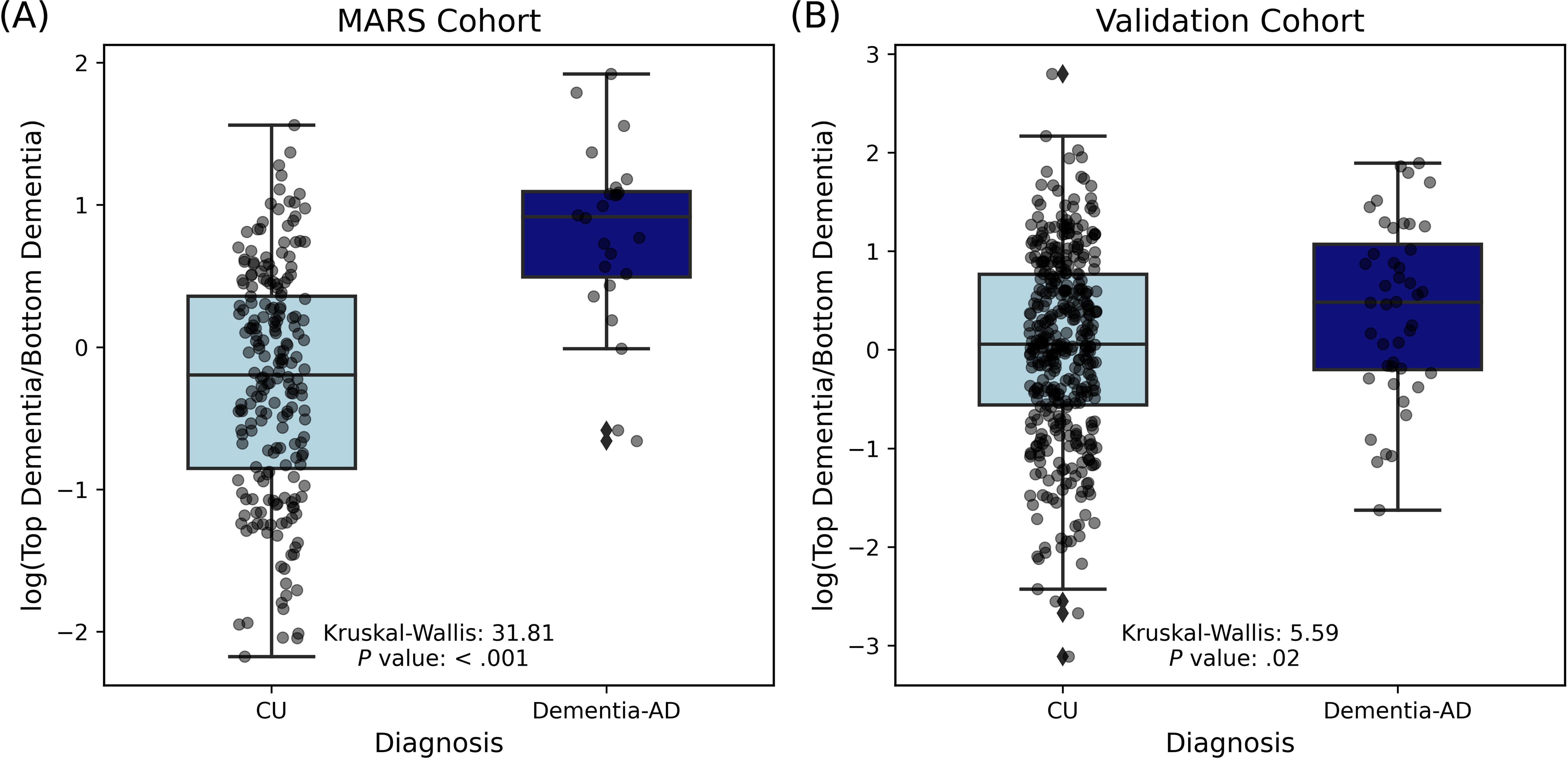
Comparison of log-transformed dementia biomarker ratios in CU and AD dementia across two cohorts. (A) The results from the MARS cohort. Box plots show the distribution of log-transformed ratios of top dementia biomarkers to bottom dementia biomarkers for CU individuals (light blue) and AD dementia (dark blue) (Kruskal-Wallis = 31.81, *P* value < .001). (B) The results from a larger validation cohort (n = 448). Box plots present the distribution of log-transformed ratios of top dementia biomarkers to bottom dementia biomarkers found in the MARS cohort for CU individuals (light blue) and people with AD dementia (dark blue) (Kruskal-Wallis = 5.59, *P* value = .02). Each point represents an individual sample, with the boxes indicating the interquartile range (IQR) and the whiskers extending to 1.5 times the IQR. The horizontal line within each box denotes the median value.

### 3.6 Gut microbiome functional pathways in a clinical diagnosis group

The DA analysis of gut microbiome functional pathways, stratified by species within each pathway, identified 116 distinct pathways that differ between individuals with AD and CU individuals (**Table S3**). Among 116 distinct pathways, we focused our analysis on pathways that only showed abundance in either the Top (more abundant in AD) or the Bottom (less abundant in AD) group. Among pathway features only with either the Top (15 pathways) or the Bottom (6 pathways) group, the log ratios of Top/Bottom features were shown to be significantly different between AD and CU (**Figure 6A**). Furthermore, gut microbiome taxonomic features that were associated with pathway features (36 features) only with either the Top or Bottom group were visualized with each pathway category (**Figure 6B**). For example, a pathway, naphthalene degradation, was one of the Bottom pathways, and microbes associated with this pathway were species *Turicibacter sanguinis*, *Bifidobacterium angulatum*, and *Lactococcus lactis* (**Figure 6B**). Another example in the Top pathway is benzoate degradation which is associated with microbes including *Anaerostipes caccae*, *Bacteroides finegoldii*, and *Bacteroides thetaiotaomicron* (**Figure 6B**).

**Figure 6.**
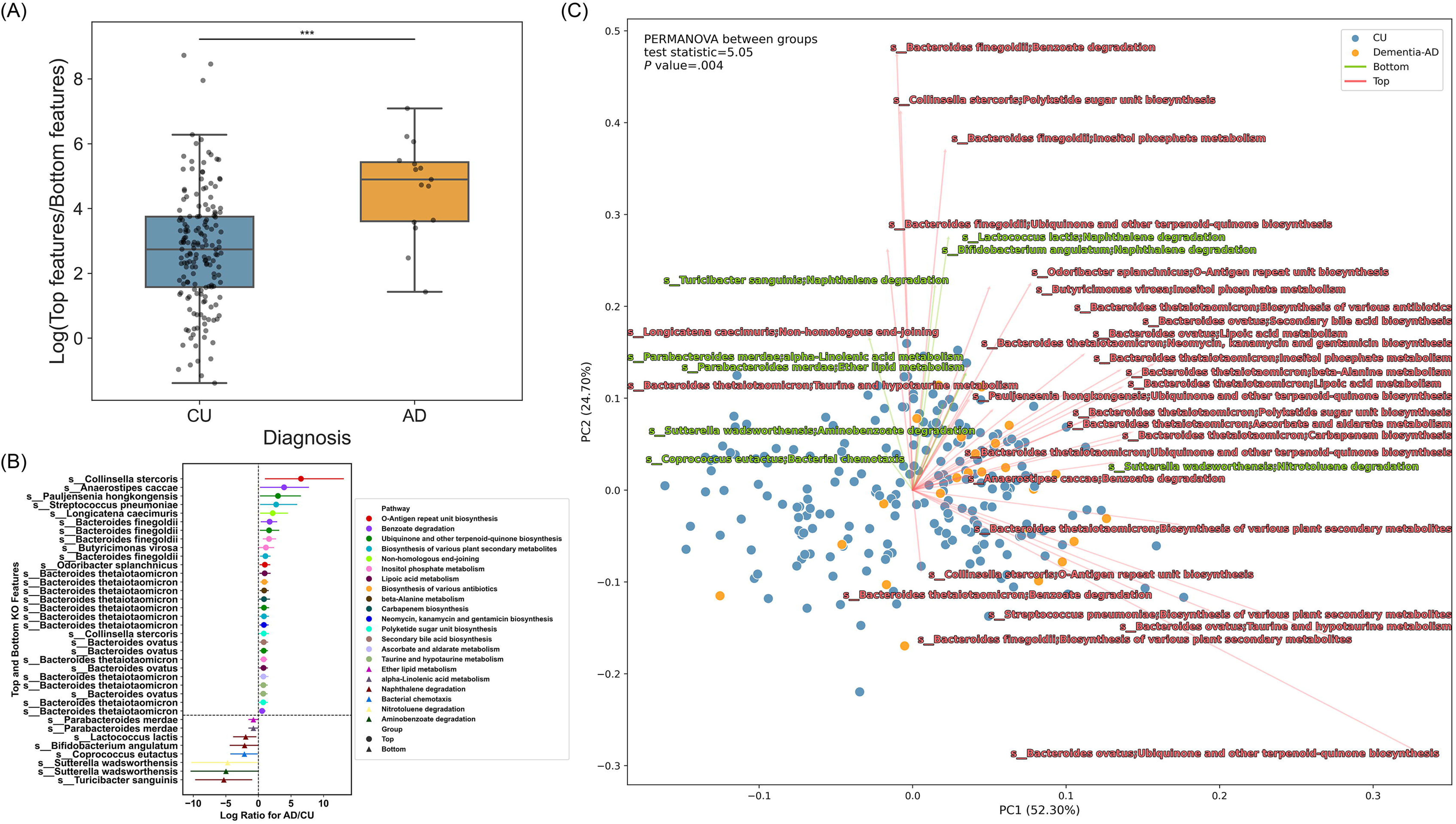
Differentially abundant microbial pathways between AD and CU. (A) The distribution of the log ratios of Top/Bottom pathway features between AD (orange) and CU (blue) was shown in a box plot. Mann–Whitney *U* test was performed to determine statistical significance. Asterisks indicate a significant difference between AD (4.90) and CU (2.74) groups in the median of the log ratios of Top/Bottom pathway features (*P* value < .001). (B) A total of 36 differentially abundant features of microbial species and their corresponding pathways between AD and CU were displayed in a forest plot. Circles denote “Top” features, indicating a positive association with AD, whereas triangles denote “Bottom” features, indicating a negative association. The lines are color-coded by unique species and their corresponding pathways. DA analysis was conducted using BIRDMAn. (C) RPCA on the clinical diagnosis group and a biplot of microbiome pathway features and their corresponding species. Each point represents an individual sample color-coded by the respective group, with CU colored in blue and AD colored in orange. Vectors represent the direction (arrows) and magnitude (length) of the contribution of feature variables to the principal components (PCs). Vectors in red indicate Top features and vectors in green indicate Bottom features. PC1 and PC2 axes represent the most variance in data. Statistical analysis on RPCA was performed with PERMANOVA between AD and CU groups.

RPCA on microbiome pathway features (36 features from DA analysis) visualized with a biplot indicated a significant separation between the clinical diagnosis group (AD and CU, *P* value = .004) (**Figure 6C**). Microbial features that belonged to the Top group indicated by the red vector directed towards many AD subjects indicated by the orange dots. Microbial features that belonged to the Bottom group indicated by the green vector directed towards many CU subjects indicated by the blue dots. For instance, multiple pathways from species *Bacteroides thetaiotaomicron* pointed towards the AD group suggesting a potentially stronger association between the Top features and AD group (**Figure 6C**). On the other hand, pathways from species *Turicibacter sanguinis*, *Bifidobacterium angulatum*, and *Lactococcus lactis*, which belonged to the Bottom group, pointed towards the CU group or showed different directions compared to the Top features suggesting a potentially weaker association between the Bottom features and AD group (**Figure 6C**).

### 3.7 Associations between gut microbiome compositional and functional features in clinical diagnosis group and CSF biomarkers of AD and related pathologies

Associations between gut microbiome features (composition and function) and CSF biomarkers of AD and related pathologies were performed as described in the ‘Statistical analysis’ section of the ‘Methods’ (**Figure 7**).

**Figure 7.**
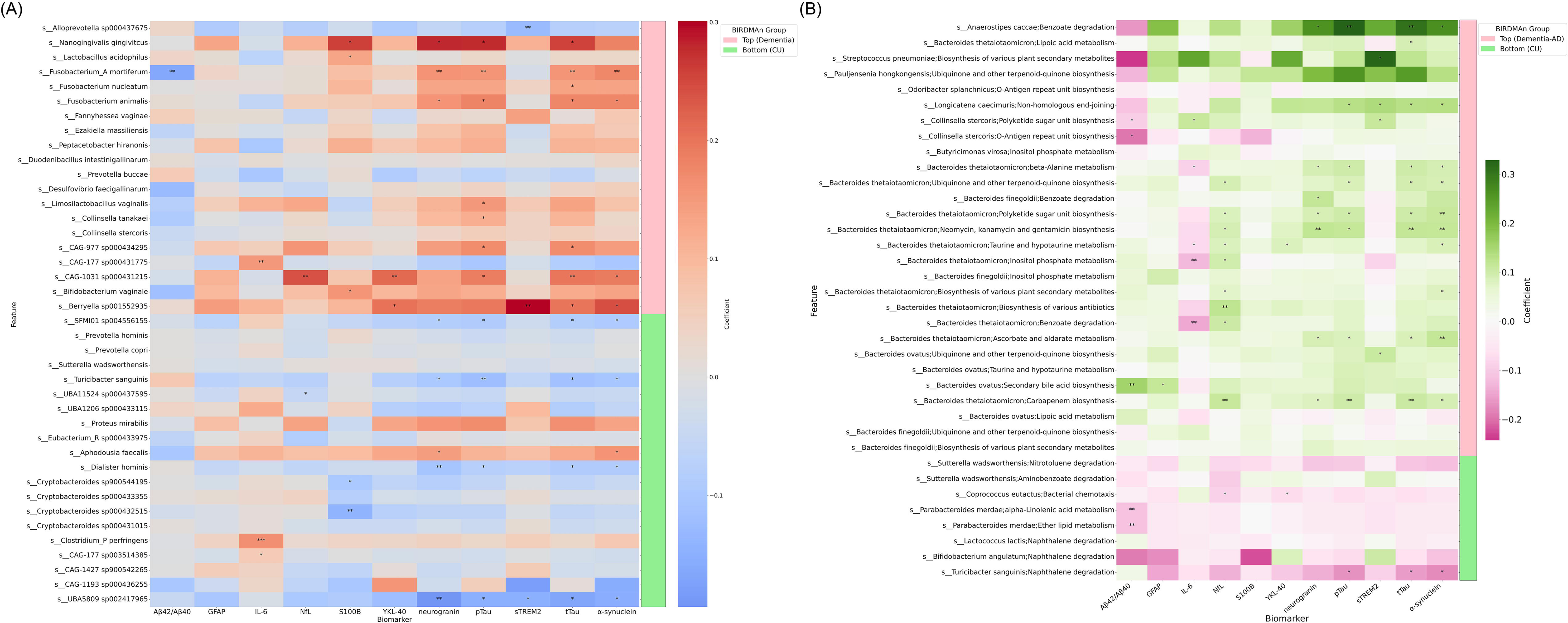
Heatmap illustrating the associations between gut microbiome compositional and functional features and CSF biomarkers in AD and related pathologies. (A) This heatmap represents the coefficients of regression analysis between the top and bottom 20 gut microbial species linked to dementia and CSF biomarkers in two groups: Top (more abundant in AD, denoted by the pink bar) and Bottom (less abundant in AD, denoted by the green bar). The color scale indicates the strength and direction of the associations, with red representing positive associations and blue representing negative associations. The intensity of the color corresponds to the magnitude of the coefficient. Listed on the left are the gut microbiome species that were identified as more or less abundant in dementia-AD through BIRDMAn. (B) The heatmap depicts the coefficients of regression analysis between the gut microbial pathways and CSF biomarkers. Coefficients are scaled by colors indicating the strength and direction of the associations, with green representing positive associations and pink representing negative associations. The intensity of the color corresponds to the magnitude (strength) of the coefficient. Microbial species and their associated pathway features are listed on the left of the plot and two groups (Top: more abundant in AD, denoted by the light pink bar; and Bottom: less abundant in AD or more abundant in CU, denoted by the light green bar) from DA analysis using BIRDMAn are displayed on the right of the plot. The biomarkers listed along the bottom include amyloid pathology (Aβ_42_/Aβ_40_), tau pathophysiology (pTau_181_ and tTau), neurodegeneration (NfL), synaptic dysfunction and injury (neurogranin and α-synuclein), inflammation (IL-6), and glial activation (S100B, GFAP, YKL-40, and sTREM2). Asterisks indicate the level of statistical significance of the associations: \*\*\**P* < .001, \*\**P* < .01, and \**P* < .05 (uncorrected).

Overall, in the association between gut microbiome compositional features and CSF biomarkers of AD and related pathologies, most species that were more abundant in AD compared to CU individuals were positively correlated with CSF biomarkers. Conversely, species that were less abundant in AD were generally negatively associated with CSF biomarkers. CSF biomarkers for AD and related pathologies included in the analysis were amyloid pathology (Aβ_42_/Aβ_40_), tau pathophysiology (pTau_181_ and tTau), neurodegeneration (NfL), synaptic dysfunction and injury (neurogranin and α-synuclein), inflammation (IL-6), and glial activation (S100B, GFAP, YKL-40, and sTREM2) (**Figure 7A**).

Species that were more abundant in AD were generally positively associated with CSF biomarkers. For example, *Nanogingivalis gingivitcus*, more abundant in AD, was positively associated with S100B, neurogranin, pTau_181_, and tTau. *Fusobacterium_A mortiferum* was positively correlated with neurogranin, pTau_181_, tTau, and α-synuclein, and negatively associated with CSF amyloid (Aβ_42_/Aβ_40_). *Fusobacterium animalis* was positively associated with neurogranin, pTau_181_, tTau, and α-synuclein. *CAG-1031 sp000431215*, a species within the Bacteroidetes phylum, was positively correlated with NfL, YKL-40, pTau_181_, tTau, and α-synuclein. *Berrvella sp001552935* was positively associated with YKL-40, sTREM2, tTau, and α-synuclein. Additionally, *Lactobacillus acidophilus* and *Bifidobacterium vaginale* were positively associated with S100B, *CAG-977 sp000434295* was associated with pTau_181_ and tTau, *Fusobacterium nucleatum* was associated with tTau, *Limosilactobacillus vaginalis* and *Collinsella tanakaei* were associated with pTau_181_, and *CAG-177 sp000431775* was associated with IL-6.

Species that were less abundant in AD were generally negatively associated with CSF biomarkers. Species *SFMI01 sp004556155*, *Turicibacter sanquinis*, and *Dialister hominis*, all within the Firmicutes phylum, were negatively associated with neurogranin, pTau_181_, tTau, and α-synuclein. *UBA5809 sp002417965*, another Firmicutes species, was also negatively associated with neurogranin, pTau_181_, tTau, and α-synuclein, as well as sTREM2. *UBA11524 sp000437595*, another Firmicutes species, was negatively associated with NfL. *Cryptobacteroides sp900544195* and *Cryptobacteroides sp000432515*, both species under phylum Bacteroidetes, were negatively associated with S100B.

In the association between gut microbiome functional features and CSF biomarkers of AD and related pathologies, multiple microbial pathways more abundant in AD compared to CU showed a tendency to positively correlate with the CSF biomarkers, whereas pathways less abundant in AD compared to CU showed a tendency to have negative associations with the CSF biomarkers (**Figure 7B**). It should be noted that a lower Aβ_42_/Aβ_40_ ratio is associated with a higher risk of having AD pathology whereas higher levels of the rest of the CSF biomarkers are associated with a higher risk of having AD pathology. The same categories of CSF biomarkers for AD and related pathologies were included in the analysis including Aβ_42_/Aβ_40_, pTau_181_, tTau, IL-6, NfL, neurogranin, α-synuclein, S100B, GFAP, YKL-40, and sTREM2.

Microbial functional features in the Top group showed overall positive associations with CSF biomarkers with the exception of Aβ_42_/Aβ_40_. Multiple *Bacteroides* spp. and their related pathways were positively associated with several CSF biomarkers including NfL, neurogranin, α-synuclein, pTau_181_, and tTau. *Bacteroides thetaiotaomicron* and its associated pathways including benzoate degradation, ubiquinone and other terpenoid-quinone biosynthesis, biosynthesis of various plant secondary metabolites, inositol phosphate metabolism, lipoic acid metabolism, biosynthesis of various antibiotics, beta-alanine metabolism, carbapenem biosynthesis, neomycin, kanamycin and gentamicin biosynthesis, polyketide sugar unit biosynthesis, ascorbate and aldarate metabolism, and taurine and hypotaurine metabolism had generally positive relationship with CSF biomarkers including NfL, YKL-40, neurogranin, α-synuclein, pTau_181_, and tTau, and negative relationship with IL-6. *Collinsella stercoris* and polyketide sugar unit biosynthesis pathway showed positive correlation with GFAP and sTREM2, and negative correlation with Aβ_42_/Aβ_40_. *Collinsella stercoris* and O-antigen repeat unit biosynthesis pathway showed negative correlation with Aβ_42_/Aβ_40_.

Microbial functional features in the Bottom group showed overall negative associations with CSF biomarkers for AD and related pathologies. Two pathways, ether lipid metabolism and alpha-linolenic acid metabolism, related to *Parabacteroides merdae*, a species more abundant in CU group were associated with lower CSF Aβ_42_/Aβ_40_. It implies that higher abundance of these pathways of *Parabacteroides merdae* in CU individuals is associated with more brain amyloid. Moreover, bacterial chemotaxis pathway from *Coprococcus eutactus* was negatively associated with NfL and YKL-40, and naphthalene degradation pathway from *Turicibacter sanguinis* was negatively associated with α-synuclein, pTau_181_, and tTau.

## 4 DISCUSSION

In this study, we compared gut microbiome composition and function between several AD-relevant groups, including those with a clinical diagnosis, differential amyloid status, and *APOE* ε*4* carrier status. The objective was to determine the association of gut microbiome features that are differentially abundant in AD dementia and determine association with CSF biomarkers of AD and related pathological features, to potentially identify gut microbial features associated with AD.

Alpha and beta diversity analysis was performed between groups of each clinical diagnosis, amyloid status, and *APOE* ε*4* status groups. Prior studies have found that alpha and beta diversities do not differ between AD vs CU^61,62^ and A+ vs A−^63^ while other studies showed significant differences in alpha and beta diversity indices in humans^4,7^ and mice.^64–66^

The DA analysis in gut microbiome composition at each taxonomic level using BIRDMAn was performed in AD-related groups. Similar results were reported in other studies^6,67^ while opposite findings were also found, where fewer Bacteroidetes and more Firmicutes were reported in MCI compared to healthy controls and fewer genera *Bacteroides* and *Alistipes* and more genus *Bifidobacterium* were found in AD compared to health controls.^68,69^ A meta-analysis of gut microbiome compositional differences in AD across studies between 2000 to 2021 demonstrated similar outcomes measured by overall pooled effect size at each taxonomic level.^7^

Discrepancies between studies may be due to differences in sample size, population variation, disease heterogeneity, sequencing method, and confounding factors.^70^ Our study addresses these discrepancies through robust methodologies. Firstly, we utilized shotgun metagenomic sequencing, which provides more comprehensive taxonomic and functional profiling of microbial communities compared to 16S rRNA sequencing.^71^ Additionally, we employed advanced statistical methodologies that have been shown to be replicable across multiple cohorts.^52^ Lastly, we validated our findings with a larger cohort from the AGMP, which significantly enhances the reliability and generalizability of our result. Our validation of the DA analysis in a larger cohort largely recapitulates what we found in the smaller sample, confirming that alterations in gut microbiome composition are present in AD dementia.

To further determine co-occurring gut microbes among clinical diagnosis, amyloid status, and *APOE* ε*4* status groups, a co-occurrence analysis was performed on differentially abundant microbes in each group. Investigating co-occurring taxonomic features may be useful in the examination of gut microbiota that could potentially coexist and contribute to AD pathology related to amyloid pathology or *APOE* ε*4* pathology which are phenotypic and genotypic pathological signatures of AD. Studies have shown gut microbiota differences between CU+(Aβ-positive) and CU− (Aβ-negative).^8,11,72^ One of the studies showed that Phylum Bacteroidetes, class Bacteroidia, and order Bacteroidales were enriched in CN+ and phylum Firmicutes, class Clostridia, order Clostridiales, families *Lachnospiraceae* and *Ruminococcaceae*, and genera *Faecalibacterium* and *Bilophila* were enriched in CN−.^11^ Higher abundance of genera *Faecalibacterium* and *Bilophila* was negatively correlated with the global brain Aβ burden.^11^ Another study explored gut microbiome taxa which are pro-inflammatory with blood inflammation markers.^73^ Genus *Escherichia*/*Shigella* was significantly more abundant in A+ compared with A− individuals. Genus *Escherichia*/*Shigella* was correlated positively with peripheral inflammatory cytokines in individuals with cognitive impairment and brain amyloidosis.^73^

Taken together, results from our study and other studies suggest that diverse and distinct gut microbiota taxonomic composition is altered in AD dementia, among individuals with preclinical AD, and individuals with genetic risk for AD. However, studies are limited to taxonomic and compositional associations and the determination of microbial functions in AD pathogenesis is needed to better understand the role of specific gut microbes and their functions in the progression of AD.

This study further examined the functional pathways of gut microbiome in a clinical diagnosis group between AD and CU. Gut microbiome pathways and associated species that are differentially abundant between AD and CU were determined. We found 116 distinct pathways between AD and CU. Among 116 KO pathway features, 21 pathways had associations either with AD (Top) or CU (Bottom) group. The log ratios of these Top/Bottom microbial pathway features between AD and CU were significantly different. Interestingly, species that were associated with these microbial pathways were mostly *Bacteroides* spp. (*B*. *finegoldii*, *B*. *thetaiotaomicron*, and *B*. *ovatus*) under phlyum Bacteroidota in the Top group.

Multiple studies have reported the association between *Bacteroides* and AD.^4,74,75^ Administration of *Bacteroides fragilis* to AD mice increased Aβ plaques and inhibition of microglial clearance of Aβ was observed after introduction to *B*. *fragilis*.^76^ Another study showed the role of *B*. *fragilis* in AD pathology in mice.^77^ Studies have suggested that genus *Bacteroides* to be dominant in older adults compared with healthy and younger controls.^78,79^ However, contrasting findings have been reported related to *Bacteroides*^8,80^ and further strain-specific studies are needed to understand the role of *Bacteroides* in AD pathology.

Additionally, RPCA showed a distinct significant separation between AD and CU groups. RPCA is known to handle sparse and high-dimensional datasets and is sensitive to datasets with outliers.^54^ Microbiome datasets are often sparse and zero-inflated, thus we employed RPCA to identify microbiome features that could explain the separation between groups. Consistent with the DA analysis, RPCA showed microbial pathways that are more abundant in AD (Top) were more associated with AD dementia, whereas microbial pathways less abundant in AD (Bottom) were less associated with AD dementia.

To determine whether distinct microbial features in AD and CU correlate with CSF biomarkers of AD and related pathologies, the relationship between CSF biomarkers and each compositional and functional gut microbiome feature was explored using the OLS regression model.

Key microbiome species, particularly *Fusobacterium nucleatum*, *Fusobacterium animalis*, and *Nanogingivalis gingivitcus*, identified as commonly more abundant between AD and A+, were associated with more intense tau pathophysiology (pTau_181_ and tTau) and/or synaptic dysfunction and injury (neurogranin and α-synuclein). Interestingly, the species *Fusobacterium nucleatum* is an oral bacteria often associated with cavity and periodontal diseases as well as colorectal cancer.^81,82^ *Fusobacterium nucleatum* produces lipopolysaccharides (LPS) that induce microglial activation with elevated expression of proinflammatory cytokines.^83^ In prior studies using the 5XFAD mouse model, the mRNA expression levels of the same proinflammatory cytokines as well as numbers of microglia in the mice brain were increased after *Fusobacterium nucleatum* infection.^83^ Moreover, enhanced Aβ accumulation, tau protein phosphorylation, and memory impairment were observed in 5XFAD mice compared to controls.^83^ The oral infection of *Fusobacterium nucleatum* in AD-like periodontitis rats exhibited increased accumulation of Aβ and pTau_181_ expression in the brain.^84^ Although these results propose valuable mechanistic backgrounds for *Fusobacterium nucleatum* and AD, further investigation in humans is needed.

The species *Dialister hominis,* a microbe that co-occurred between CU and A− displayed a negative correlation with AD pathology (pTau_181_), neuronal damage (tTau), and synaptic dysfunction and injury (neurogranin and α-synuclein). These findings are similar to previous works which identified genus *Dialister* (less in AD) to be more abundant in CU individuals.^85,86^ Another species *Turicibacter sanguinis* which was a microbe that co-occurred between CU and *APOE* ε*4*− showed a negative correlation with pTau_181_, tTau, neurogranin, and α-synuclein. In animal models for AD, *Turicibacter sanguinis* is reported to be less abundant in AD compared to controls.^87,88^ In humans, *Turicibacter* was observed to be less abundant in individuals with AD.^4^ However, studies on the role of species specific to *Dialister hominis* and *Turicibacter sanguinis* in AD pathology are scarce.

This result indicates that microbiota compositional features in CU are related to lower levels of AD pathology whereas microbiota features in AD are related to higher levels of AD pathology, suggesting overall microbiota composition in people with AD may be vulnerable to development or progression of AD compared to CU individuals. While these results support a relationship between gut microbiome composition and AD pathology, further investigation into these mechanisms is required to find a causal relationship.

Multiple *Bacteroides* spp. and their related functional pathways more abundant in AD were associated with greater AD pathology represented in CSF biomarkers of AD and related pathologies. Biomarkers including neurodegeneration (NfL), synaptic dysfunction and injury (neurogranin and α-synuclein), and tau pathophysiology (pTau_181_ and tTau) showed a positive relationship with *Bacteroides thetaiotaomicron* and their functions. Studies have linked the abundance of *Bacteroides thetaiotaomicron* with AD. The abundance of *B*. *thetaiotaomicron* was significantly higher in AD mice and was related to poorer spatial learning.^89^ Increased abundance of *B*. *thetaiotaomicron* was reported in AD participants.^90,91^ However, in a non-AD model, *B*. *thetaiotaomicron* was suggested to regulate enteric neuronal cell populations and neurogenic function.

Although evidence related to these microbial functions is limited, these results suggest that alterations in gut microbiome composition and function are related to AD pathological markers measured in CSF.

The main limitation of our study is the small number of cognitively impaired participants relative to CU participants. The sample size decreased after matching clinical measurements, gut microbiome data, and presence of CSF biomarkers. The resulting low statistical power may have led to losing significance after multiple test corrections for association analyses between gut microbiome features and CSF biomarkers. A similar challenge is the inclusion of cognitively impaired individuals in both the A+ and *APOE* ε*4* groups. Excluding these individuals reduced the sample sizes, resulting in low statistical power. Future studies should aim to collect sufficient samples to separate these groups, allowing for a clearer distinction between disease effects and symptom effects. Moreover, due to the cross-sectional approach, it is difficult to capture the longitudinal changes over time considering the progression of AD for each individual. We included the ADRC validation cohort to account for differences in gut microbiome composition across diverse populations, however, due to the limitation of the availability of biomarkers matched with fecal samples, we were not able to test associations with CSF biomarkers in the validation cohort. Additionally, the results are correlational, and further mechanistic studies are needed to find causal relationships between gut microbiome features and biomarkers for AD pathology. Finally, other environmental factors (exposome) which may impact the gut microbiome and which contribute to AD risk require additional study in the future.

## 5 CONCLUSIONS

This study suggests that gut microbiome composition and function differ between people with AD dementia and CU individuals. Beta diversity indices differed among AD-related groups: diagnosis (AD vs CU), amyloid (A+ vs A−), and *APOE* ε*4* (*APOE* ε*4*+ vs *APOE* ε*4*−) groups, indicating that the gut microbiome diversity varies between each group. Multiple gut microbes at each taxonomic level including phylum, family, genus, and species were differentially abundant across AD groups. Co-occurring gut microbes across AD-related groups were determined, many of which showed associations with CSF biomarkers for AD and related pathologies. Microbial functional pathways were differentially abundant between AD and CU, which were correlated with AD pathology markers measured in CSF. These findings identify specific targets for stratifying key gut microbes and microbial pathways that may be related to AD pathology. Further investigation on metabolomic changes as well as exposome and host genome that may be mediating the interconnectome between the gut microbiome and AD pathology is needed.

## ACKNOWLEDGMENTS

This project was enabled in part by The Alzheimer Gut Microbiome Project (AGMP) and the Alzheimer’s Disease Metabolomics Consortium (ADMC) funded wholly or in part by the following NIA grants thereto: U01AG061359 and U19AG063744 awarded to Dr. Kaddurah-Daouk at Duke University in partnership with multiple academic institutions. As such, the investigators within the AGMP and the ADMC, not listed specifically in this publication’s author’s list, provided data along with its pre-processing and prepared it for analysis, but did not participate in analysis or writing of this manuscript. A listing of AGMP Investigators can be found at alzheimergut.org/meet-the-team/. A complete listing of ADMC investigators can be found at: sites.duke.edu/adnimetab/team/. The IGM S10 grant S10 OD026929 was awarded to Dr. Rob Knight at the University of California San Diego IGM Genomics Center. This publication includes data generated at the University of California San Diego IGM Genomics Center utilizing an Illumina NovaSeq 6000 that was purchased with funding from a National Institutes of Health SIG grant (#S10 OD026929).

This work was also supported by the National Institute on Aging Grant R01AG070973 (B.B.B., F.E.R., T.K.U.), National Institute on Aging Grant R01AG083883 (T.K.U., B.B.B., F.E.R.), Vilas Early-Career Investigator Award (T.K.U.), and National Institute on Aging Grant P30AG062715 (which in part supported B.B.B., T.K.U.).

Dr. Zetterberg is a Wallenberg Scholar and a Distinguished Professor at the Swedish Research Council supported by grants from the Swedish Research Council (#2023-00356; #2022-01018 and #2019-02397), the European Union’s Horizon Europe research and innovation programme under grant agreement No 101053962, Swedish State Support for Clinical Research (#ALFGBG-71320), the Alzheimer Drug Discovery Foundation (ADDF), USA (#201809-2016862), the AD Strategic Fund and the Alzheimer’s Association (#ADSF-21-831376-C, #ADSF-21-831381-C, #ADSF-21-831377-C, and #ADSF-24-1284328-C), the European Partnership on Metrology, co-financed from the European Union’s Horizon Europe Research and Innovation Programme and by the Participating States (NEuroBioStand, #22HLT07), the Bluefield Project, Cure Alzheimer’s Fund, the Olav Thon Foundation, the Erling-Persson Family Foundation, Familjen Rönströms Stiftelse, Stiftelsen för Gamla Tjänarinnor, Hjärnfonden, Sweden (#FO2022-0270), the European Union’s Horizon 2020 research and innovation programme under the Marie Skłodowska-Curie grant agreement No 860197 (MIRIADE), the European Union Joint Programme – Neurodegenerative Disease Research (JPND2021-00694), the National Institute for Health and Care Research University College London Hospitals Biomedical Research Centre, and the UK Dementia Research Institute at UCL (UKDRI-1003). COBAS is a trademark of Roche. All other product names and trademarks are the property of their respective owners. The NeuroToolKit is a panel of exploratory prototype assays designed to robustly evaluate biomarkers associated with key pathologic events characteristic of AD and other neurological disorders, used for research purposes only and not approved for clinical use (Roche Diagnostics International Ltd, Rotkreuz, Switzerland).

## CONSENT STATEMENT

All human subjects provided informed consent to participate in this study.

## DATA AVAILABILITY

Samples were provided by the University of Wisconsin Alzheimer’s Disease Research Center. Clinical data can be requested from the National Alzheimer’s Coordinating Center (naccdata.org/).

Data will be available in the Synapse AD Knowledge Portal.

Gut Microbiome data is stored and accessible via the University of California, San Diego Qiita platform (qiita.ucsd.edu/).

