## Supplementary material for "Gut microbiome compositional and functional features associate with Alzheimer’s disease pathology": Table S1

**Table S1. Validation cohort demographics at fecal sample collection by clinical diagnosis.**

| **Variable** | **N** | **Overall**, N = 448*^†^* | **Dementia-AD**, N = 44*^†^* | **CU**, N = 404*^†^* |
| --- | --- | --- | --- | --- |
| **Age** | 448 | 72 (±8) | 73 (±9)*^‡**^* | 72 (±8) |
| **Sex** | 448 |  |  |  |
| Female |  | 285 (64%) | 22 (50%) | 263 (65%) |
| Male |  | 163 (36%) | 22 (50%) | 141 (35%) |
| **Race** | 433 |  |  |  |
| Black or African American |  | 50 (12%) | 1 (0.02%) | 49 (12%) |
| White |  | 383 (88%) | 40 (98%) | 343 (88%) |
| ***APOE* genotype** | 416 |  |  |  |
| *ε*2*ε*3 |  | 48 (12%) | 3 (9%) | 45 (12%) |
| *ε*3*ε*3 |  | 217 (52%) | 14 (41%)*^‡**^* | 203 (53%) |
| *ε*2*ε*4 |  | 13 (3%) | 1 (3%) | 12 (3%) |
| *ε*3*ε*4 |  | 118 (28%) | 12 (35%) | 106 (28%) |
| *ε*4*ε*4 |  | 20 (5%) | 4 (12%)*^‡****^* | 16 (4%) |
| ***APOE ε4* genotype** | 416 |  |  |  |
| Negative (non-carrier) |  | 265 (64%) | 17 (50%) | 248 (65%) |
| Positive (carrier) |  | 151 (36%) | 17 (50%)*^‡****^* | 134 (35%) |
| **BMI** | 315 | 27 (±5) | 25 (±4)*^‡*^* | 27 (±5) |
| **Amyloid status** | 65 |  |  |  |
| 0 (A−) |  | 45 (69%) | 1 (12%) | 44 (77%) |
| 1 (A+) |  | 20 (31%) | 7 (88%)*^‡****^* | 13 (23%) |
| Abbreviations: AD, Alzheimer’s disease; CU, cognitively unimpaired; *APOE*, apolipoprotein E; BMI, body mass index; A, amyloid status. | | | | |
| *^†^*Mean (±SD); n (%) | | | | |
| *^‡^*Significantly different Dementia-AD vs CU | | | | |
| **P* < .05, ***P* < .01, ****P* < .001, *****P* < .0001 (*P* values are Bonferroni test corrected) | | | | |
